# Causal effects of body mass index on glycaemic traits in African and European populations: a Mendelian randomization study

**DOI:** 10.64898/2026.10.02.26364581

**Authors:** Raylton P Chikwati, Jean-Tristan Brandenburg, Dhriti Sengupta, Michele Ramsay, Nigel J Crowther, Ana L Goncalves Soares, Amanda HW Chong, Deborah A Lawlor

## Abstract

**Aims/hypothesis:** Higher body mass index (BMI) is associated with higher levels of HbA1c and glucose in conventional observational studies from sub-Saharan Africa, but its causal relationship in this population has not been explored. Mendelian randomization (MR) which leverages genetic variants can strengthen causal inference. However, its application in African ancestry populations is limited. The aim of this study was to determine whether causal effects of BMI on glycated haemoglobin (HbA1c) and random glucose differ between African and European populations.

**Methods:** We used two-sample MR to estimate the effects of BMI on HbA1c and random glucose in adults of African and European ancestries. We conducted a cross-ancestry BMI genome-wide association study (GWAS) from the Africa-Wits International Network for the Demographic Evaluation of Populations and Their Health (INDEPTH) Partnership for Genomic Studies (AWI-Gen) dataset of African adults (N=10,476) and the largest BMI GWAS in Europeans (N∼700,000). From the cross-ancestry GWAS we identified 428 independent genome-wide significant variants and of those, 363 single nucleotide polymorphisms (SNPs) had consistent direction of effect in both Africans and Europeans. These variants were then used as genetic instruments to proxy BMI for both Africans and Europeans. Primary outcome data for African and European populations were obtained from UK Biobank summary statistics for HbA1c (5,790 Africans and 400,825 Europeans) and random glucose (5,750 Africans and 314,916 Europeans), with additional results for HbA1c from the MAGIC multi-ancestry GWAS comprised of Ugandan individuals (N=4,441), African Americans (N= 6,647), and Europeans (N=146,806). Main analyses used the inverse variance weighted (IVW) method, with sensitivity analyses conducted using MR-Egger, weighed median and weighted mode approaches.

**Results:** The main IVW analyses in UK Biobank provided support for higher BMI causally increasing HbA1c levels in both Africans (0.26 SD higher HbA1c per 1 SD BMI, 95% CI 0.07, 0.45) and Europeans (0.27 SD higher HbA1c per 1 SD BMI, 95% CI 0.22, 0.32). UK Biobank main analyses also supported higher BMI increasing random glucose in Africans (0.21 SD higher mean glucose per 1 SD BMI, 95%CI 0.03, 0.37) and Europeans (0.15 SD higher mean glucose per 1 SD BMI, 95%CI 0.12, 0.18). Sensitivity analyses suggested these results were not biased by unbalanced horizontal pleiotropy. Results from MAGIC also showed that higher BMI causally increased HbA1c in Europeans (0.04% higher HbA1c per 1 SD BMI, 95% CI 0.02, 0.05), though results for African Americans (0.01% higher HbA1c per 1 SD BMI, 95% CI -0.05, 0.07) and Ugandans (0.02% lower HbA1c per 1 SD BMI, 95% CI -0.10, 0.05) spanned the null.

**Conclusion/interpretation:** We have found broadly similar effects of BMI on HbA1c and random blood glucose in African and European populations. However, much larger GWAS of glycaemic traits in African and other non-European populations are needed to strengthen causal inference and support equitable translation of these findings. The method that we have used to obtain identical strong genetic instrumental variables (IVs) for both European and African populations may have value for other MR studies interested in exploring differences in effects between different population groups.

**Research in Context:** *What is already known about this subject?:*

- Conventional observational studies in sub-Saharan Africa show a positive association between higher body mass index (BMI) and higher glycated haemoglobin (HbA1c) and glucose levels.
- These associations may be influenced by confounding and reverse causation.
- Mendelian randomization (MR) evidence in European populations show that higher BMI is causally associated with increased HbA1c and glucose, but the application of MR in African ancestry populations is limited.

*What is the key question?:*

- Does higher BMI causally influence the levels of HbA1c and random glucose similarly in African and European populations?

*What are the new findings?:*

- Higher BMI increased HbA1c and random glucose in both African and European adults, with similar estimates in both populations.
- We demonstrated how MR can be used to investigate population-specific differences in the health effects of the same exposures between African and European populations.

*How might this impact on clinical practice in the foreseeable future?:*

- Improved causal understanding may strengthen prevention strategies targeting adiposity to reduce the risk of type 2 diabetes in sub-Saharan African populations.

## Introduction

The global obesity epidemic is evident in sub-Saharan Africa, where the prevalence of adult obesity reached 12.1% in 2022, an increase of approximately 40% from 8.6% in 2010, as estimated by the World Health Organization [1]. This upward trend is likely to contribute to the increased burden of type 2 diabetes and other cardiometabolic diseases among adults in the region. Conventional observational studies in sub-Saharan Africa show a positive association between high body mass index (BMI) and diabetes [2–6], fasting glucose [7, 8], and glycated haemoglobin (HbA1c) [3]. Whether these associations are causal, and if so whether the effects of higher BMI on these outcomes in sub-Saharan African populations are the same as those in European populations is unclear.

Mendelian randomization (MR) is a method that commonly uses genetic variants (i.e., single nucleotide polymorphisms (SNPs)) as instrumental variables (IV) for the exposure of interest to obtain the unconfounded effect of that exposure on an outcome [9]. Whilst MR is less prone to confounding by socioeconomic, behavioural and health factors and reverse causality that commonly affect conventional observational studies [10], it is reliant on not violating three key assumptions: (i) the genetic IV should be statistically strongly associated with the exposure in the relevant population (relevance assumption), (ii) there should be no confounding between the genetic IV and the outcome (independence assumption), (iii) the genetic IV should only affect the outcome via the exposure of interest (exclusion restriction criteria).

Previous studies using MR in European populations have supported the causal effects of higher BMI on glycaemic traits including fasting glucose [11] and HbA1c [12], and also demonstrated that these effects on glycaemic traits mediate the effect of higher BMI on coronary heart disease [13]. The use of MR in sub-Saharan Africa remains limited due to scarce genetic data and limited local expertise. However, growing population-based studies, such as the Africa-Wits International Network for the Demographic Evaluation of Populations and Their Health (INDEPTH) Partnership for Genomic Studies (AWI-Gen) [14], present an opportunity for expanding MR studies in the region. This has potential to increase our understanding of the aetiology of cardiometabolic diseases such as type 2 diabetes, to inform prevention strategies. It is well understood that variation in allele frequencies and patterns of correlated SNPs (i.e., those in linkage disequilibrium (LD)) between populations can limit the applicability of MR studies conducted in one ancestry to inform causal effects in individuals of other ancestries [15]. Furthermore, diabetes occurs at lower BMI in sub-Saharan Africa, and we previously showed in AWI-Gen a positive but modest association between BMI and type 2 diabetes risk, suggesting BMI alone may not fully explain causation [5, 16]. MR can help elucidate the causal role of BMI in these settings.

The aim of this study was to determine the causal effect of higher BMI on random glucose and HbA1c in African populations and compare these findings to those in European populations using MR. Full details of the ascertainment of ethnicity/ancestral information are provided in the Methods.

## Methods

We used several data sources for our analyses which included AWI-Gen, GIANT (the Genetic Investigation of ANthropometric Traits), the UK Biobank and MAGIC (the Meta-Analyses of Glucose and Insulin-related traits Consortium) (Table 1).

**Table 1:** Data sources.

|  | Trait | Data source | Population, N |
| --- | --- | --- | --- |
| Exposure | BMI | AWI-Gen | Africans, 10,476 |
|  |  | GIANT | Europeans, 681,275 |
| Outcomes | HbA1c | UK Biobank | Africans, 5,790<br>Europeans, 400,825 |
|  |  | MAGIC | Africans living in Uganda, 4,441<br>African Americans, 6,647<br>Europeans, 146,806 |
|  | Random glucose | UK Biobank | Africans, 5,750<br>Europeans, 314,916 |
|  | Fasting Glucose | AWI-Gen | Africans, 11,602 |

AWI Gen: Africa Wits INDEPTH Partnership for Genomic studies. GIANT: Genetic Investigation of ANthropometric Traits (GIANT; included participants from the UK Biobank). MAGIC: Meta Analyses of Glucose and Insulin related traits Consortium.

### Evaluation of African cohorts and data

In our initial search for BMI GWAS on May 10, 2026, we searched the GWAS catalog [17] and IEU OpenGWAS repository [18] (details of the search in the Supplementary Text). Our findings identified only one GWAS undertaken in Africa [19], which combined summary data from four cohorts: Uganda Genome Resource (UGR, East Africa; n=6,407, [19]), Durban Diabetes Study (DDS, South Africa; n=1,165, [20]), Diabetes Case Control (DCC, South Africa; n=1,542, [21]), and Africa America Diabetes Mellitus (AADM, West and East Africa; n=5,231, [22]). This GWAS included a total of 14,345 participants and identified two genome-wide significant BMI loci (rs12405634 on chromosome 1 in the intergenic region between *PLD5* and *SDCCAG8* and rs7798566 on chromosome 7 in the intergenic region within the TAS2R gene family). Of the four studies included in this GWAS, one only includes participants with diabetes [21], and one is a case-control study with 44.8% of the participants having diabetes [22]. These two studies contributed 47.2% of the total participants in this GWAS. The other two studies had participants who were from the general population. Thus, this BMI GWAS has to some extent adjusted for diabetes, and we know that adjusting for a heritable trait in GWAS that are then used in two-sample MR can introduce bias [23]. In particular, as our outcomes are all strongly related to diabetes, using the results from this GWAS could produce biased results. The remaining two studies [19, 20] together included 7,572 participants. Summary data for these two studies were not publicly available, meaning we could not use them to obtain genetic variants for the IV and even if they were, the number of participants would be smaller than that in the AWI-Gen cohort (n=11,702). For these reasons we chose to use the AWI-Gen cohort as the primary cohort for our exposure of interest, in addition to GIANT.

### Study populations and data

#### AWI-Gen (Fasting glucose outcomes and BMI GWAS summary statistics-continental African)

The AWI-Gen data consists of 11,702 participants primarily aged 40-60 years, of whom 562 were older than 60 years [14]. Participants were living in one of six African centres at the time of recruitment (between 2013 and 2017): three in South Africa (Agincourt; n=1,465, DIMAMO (formerly known as Dikgale; n=1,167), and Soweto; n=2,030), one in East Africa (Nairobi, Kenya; n=1,942), and two in West Africa (Navrongo, Ghana; n=2,014, and Nanoro, Burkina Faso; n=2,084) [14]. Ethnicity was self-reported, with all participants self-identifying as belonging to an African ethnic group. Fasting glucose was measured from venous blood samples collected after an overnight fast of at least 8 hours and analysed using a Randox Plus clinical chemistry analyser (Randox Laboratories Ltd, Crumlin, UK).

A total of 11,602 DNA samples from AWI-Gen participants were genotyped using the 2.3 million H3Africa SNP array at the Illumina® FastTrackTM Microarray Services (Illumina, San Diego, USA). Genotype quality control, imputation, and post-imputation QC were performed as described previously [24, 25]. Briefly, at the variant level, SNPs with missingness >0.05, minor allele frequency (MAF) <0.01, Hardy-Weinberg equilibrium p-value <0.0001, and autosomal SNPs that did not match the GRCh37 reference were excluded. X chromosome, Y chromosome and mitochondrial SNPs were also removed. Supplementary Figure 1 illustrates the main steps and criteria through which SNPs and DNA samples were selected. At the sample level, individuals with genotype missingness >0.05, evidence of sample duplication (PI_HAT >0.9), or discordance between recorded and genetic sex were excluded. Principal component analysis (PCA) was performed using a linkage disequilibrium-pruned subset of SNPs with the smartPCA program implemented in EIGENSTRAT [27] to assess population structure.

Genotype imputation was subsequently performed using the African Genome Resources reference panel through the Sanger Imputation Server [28]. Post-imputation quality control excluded rare variants (MAF ≤0.01) and poorly imputed variants (Info score ≤0.6).

For the BMI association analysis, participants with missing BMI values (n=57) and those with extreme BMI values (>6 SD from the mean; n=2) were excluded. Individuals with extreme values on any of the first 10 principal components (>7 SD from the mean; n=67) were also excluded as genetic outliers. A total of 10,476 participants were therefore included in the final BMI GWAS (Table 1 and Supplementary Figure 1). BMI values were inverse-rank normal transformed prior to association testing. Genome-wide association analysis was performed using a linear mixed model implemented in BOLT-LMM (v2.3.4), with adjustment for age, sex, and the first 10 principal components.

#### GIANT (BMI GWAS summary statistics - European)

We used the most recent BMI GWAS for Europeans, the Genetic Investigation of ANthropometric Traits (GIANT) GWAS (2018) which included ∼700,000 adults comprising of European-ancestry participants from UK Biobank and earlier GIANT cohorts [26]. Genetic ancestry was inferred by projecting participants onto PCs derived from 1000 Genomes reference populations and assigning individuals to the nearest of five ancestry super-populations (European, African, East Asian, South Asian, and Admixed American). Only individuals classified as European ancestry were included in the GWAS. Genome-wide association testing for BMI in the UK Biobank was performed using a linear mixed model implemented in BOLT-LMM v2.3. The resulting BMI summary statistics were subsequently combined with those from earlier GIANT cohorts using fixed-effect inverse-variance weighted meta-analysis implemented in METAL, with a total of 681,275 participants included in the meta-analysis [26]. Linear models were adjusted for age, sex, recruitment centre, genotyping batches and 10 ancestry PCs. Following covariate adjustment, residuals were inverse-rank normal transformed and standardised, such that effect estimates are expressed in SD units [26].

#### UK Biobank (HbA1c and random glucose outcome GWAS summary statistics European ancestry and African ancestry)

UK Biobank data consists of 503,317 participants aged 40-69 years who were living in the UK between 2006 and 2010 and provided sociodemographic and behavioural data, as well as blood samples for HbA1c, random glucose, and genotyping data HbA1c was measured in mmol/mol by high-performance liquid chromatography on a Bio-Rad VARIANT II Turbo analyser. Random glucose was measured in participants who were not instructed to fast before attending the assessment visit (i.e. non-fasting) and was measured in mmol/L by hexokinase analysis on a Beckman Coulter AU5800. We identified GWAS for both Africans and Europeans with summary data for HbA1c (5,290 Africans and 400,825 Europeans), and random glucose (5,750 Africans and 314,916 European) [29] (Table 1). Genotyping was performed on 488,377 participants using the UK BiLEVE (N=49,979) and UK Biobank Axiom (N=438,398) arrays. African and European ancestry was defined using genetic ancestry clusters assigned to continental ancestry groups through comparison with the Haplotype Reference Consortium, UK10K, and 1000 Genomes reference panels. All GWAS data excluded participants with known diabetes. GWAS data from the UK Biobank was based on inverse normal-transformed HbA1c (yielding effect estimates in SD units).

#### MAGIC (HbA1c outcome GWAS summary statistics - European ancestry, African American and continental African ancestry)

We identified summary GWAS statistics for HBA1c from the MAGIC Consortium multi-ancestry GWAS which had separate data for three ancestral groups: African American (N= 6,647); continental African adults living in Uganda (N=4,441), and European (N=146,806) [30, 31]. There was, however, no available data on random glucose unadjusted for BMI for African and European populations. All GWAS data excluded participants with known diabetes. HbA1c measurements were obtained from participating cohorts and were assayed using cohort-specific laboratory methods. Genotyping was conducted using different genome-wide arrays or the Illumina CardioMetabochip. Association analyses were performed within each cohort and subsequently combined through trans-ancestry GWAS meta-analysis. Analyses were adjusted for study-specific covariates including age, sex, recruitment centre, and ancestry PCs following cohort-specific quality control procedures. MAGIC data was based on raw HbA1c expressed as % units.

#### Ethics

Ethical approval for the recruitment of participants in the AWI-Gen study was obtained from the Human Research Ethics Committee (Medical) of the University of the Witwatersrand (M121029 and M2210108). Local ethics approval was obtained from the respective in-country ethical bodies. All participants provided written informed consent following community engagement and individual information sharing. The MR analyses used publicly available GWAS summary statistics from studies that had received ethical approval from their respective institutions and obtained informed consent from participants. Only anonymised summary-level data were accessed; therefore, no additional ethical approval was required for these analyses.

#### BMI Instrument selection for the cross-ancestry African and European GWAS

As the GIANT BMI GWAS in European individuals N ∼700,000 [26] is considerably larger than the AWI-Gen BMI GWAS (N=10,476), it was likely that meaningful comparisons between effects in the two populations could not be made because of the marked differences in genetic IV strength. Specifically, the African IV would be weak, leading to bias towards the null, resulting in a potentially incorrect interpretation of higher BMI in Africans having a weaker association with the outcome variable than in Europeans. Therefore, we adopted a multi-ancestry GWAS by combining the AWI-Gen and GIANT GWAS datasets and explored heterogeneity in genetic associations between the two populations. While multi-ancestry GWAS approaches have been previously described [32], our application of this framework to derive population-specific IVs of comparable strength for subsequent analyses represents a novel aspect of the present study.

We first conducted SNP harmonisation using the AWI-Gen and GIANT BMI GWAS summary statistics to align the direction of the effect allele and beta effect estimates. We performed this by matching SNP IDs based on chromosome and base-pair positions. Summary statistics from the AWI-Gen and GIANT BMI GWAS were harmonised by matching variants on chromosome, base-pair position, and alleles. Variants with matching allele pairs across both datasets were retained. Effect alleles in GIANT were oriented to represent the BMI-increasing allele, where necessary, effect and non-effect allele designations were swapped, beta estimates were multiplied by minus 1, and effect allele frequencies were recalculated as 1 minus the original frequency. The AWI-Gen statistics were subsequently aligned to match the GIANT effect allele, with corresponding adjustments made to effect sizes and frequencies. This ensured a unified effect allele orientation for every variant across both cohorts.

We then compared genetic effects between the African and European populations to account for differences in LD structures across ancestries. We conducted a fixed-effect meta-analysis of the AWI-Gen and GIANT BMI GWAS using METAL [33] to combine the association estimates across the two populations. This provided a common set of variants for subsequent instrument selection and enabled the identification of cross-ancestry genetic instruments for the MR analyses. In total, 12,752,139 common SNPs were included in the meta-analysis.

Independent SNPs were identified by first choosing SNPs on the basis that they were strongly associated with BMI (based on the genome-wide significance threshold; p ≤ 5 x 10^-8^). We then grouped correlated SNPs (i.e., those in LD and defined by r^2^), and when two or more SNPs were correlated, the SNP with the strongest association with BMI was retained. The identification of these SNPs was performed on a combined African-European reference panel (10 Mb window, r^2^ < 0.001) comprising of whole genome sequence data from European populations in the 1000 Genomes Project (Phase 3) and the African populations from the AWI-Gen study which were used to estimate LD between SNPs [34]. This resulted in 428 independent SNPs.

#### Harmonisation of SNPs across African and European GWAS

For each outcome and exposure dataset, we aligned chromosome and base-pair positions to retain only matching variants and ensured that the effect allele in the outcome dataset corresponded to the same effect allele in the exposure dataset. Where orientation differed, we reversed the beta estimate and recalculated the effect allele frequency. We used the exposure effect allele frequency as a proxy for MAF and excluded misaligned SNPs with large effect allele frequency discrepancies (>0.25) between exposure and outcome datasets. Palindromic SNPs (A/T or G/C) were retained, and strand alignment was resolved using allele frequency information to ensure consistent orientation between exposure and outcome datasets (MAF >0.25).

### Statistical analyses

#### Comparison of SNP associations with BMI between African and European populations

For each of the 428 genome-wide independent significant SNPs we compared the magnitude and direction of their association with BMI between the African and European populations. For this analysis, we use the Cochran’s Q heterogeneity test, and given the limited statistical power, we used a p-value threshold of <0.1, as is common in meta-analyses, to indicate statistical evidence of heterogeneity. The relevance to each population was explored by testing heterogeneity between each population for the associations of each genome-wide significant SNP in the cross-ancestry BMI GWAS.

#### Two-sample MR analyses (main analyses)

For our main analyses we used IVW random-effects two-sample MR to estimate the causal effect of higher BMI on HbA1c, and random glucose. The IVW method pools results for each SNP by fitting a linear regression of the SNP-outcome association on the SNP-exposure association for each SNP and constraining the intercept to be zero [35]. Each SNP is weighted by the inverse of its variance to produce an overall causal estimate.

##### Sensitivity analyses and exploration of violation of assumptions

We evaluated violations to the core MR assumptions by firstly assessing instrument strength in Africans and Europeans using the mean F-statistic for the association of each IV with BMI.

To explore bias due to unbalanced horizontal pleiotropy we compared the main IVW results to those from three methods that are more robust to pleiotropy: MR-Egger, weighted median, and weighted mode. The MR-Egger method is the same as the IVW except it does not constrain the intercept to be zero. Instead, the intercept reflects the best fitting regression, meaning that the effect estimate is corrected for bias due to unbalanced horizontal pleiotropy and the p-value for the intercept is a statistical test of the presence of unbalanced horizontal pleiotropy in the main IVW result [36]. The weighted median result is valid if at least 50% of the weight of genetic instruments are not affected by horizontal pleiotropy [37]. The weighted mode result assumes that the most common (modal) causal effect estimate, weighted by instrument strength, corresponds to the largest group of valid SNPs with no unbalanced horizontal pleiotropy [38].

#### Exploratory one-sample MR analyses

We undertook exploratory one-sample MR analyses using the AWI-Gen data. This enabled us to explore potential effects in Africans living in Africa, with both the genetic IV and outcome data in the same population. Whilst this provides relevant data for an African population, it is limited in terms of statistical power, ability to test bias due to unbalanced horizontal pleiotropy, and lack of data on our two main outcomes of interest i.e., HbA1c and random glucose. For these reasons we considered these analyses to be exploratory. The only glycaemic trait in AWI-Gen is fasting glucose, and we assumed directional consistency in the results for fasting glucose in the AWI-Gen one sample MR with the main two sample MR for Africans being supportive of those main results. We included 10,476 participants who had genotype and fasting glucose data. We used the same BMI genetic variants used in the two-sample MR as IVs for BMI. We generated a weighted genetic risk score (GRS) for BMI for each participant. This was done by counting the number of alleles positively associated with each SNP i.e., 0, 1, or 2, and multiplying that count by the corresponding β-coefficient of the SNP with BMI and then summing the weighted values for each SNP. We then analysed the association of the GRS with fasting glucose levels (inverse-transformed; mmol/L) using multivariable linear regression, adjusting for age, sex, study centre and five ancestry PCs. As with two-sample MR, between-SNP heterogeneity could indicate violation of any of the three key assumptions, and we explored this using Cochran’s Q test. Instrument strength was assessed using the F-statistic for the association of the weighted GRS with BMI in the AWI-Gen population. Between-SNP heterogeneity was assessed using Cochran’s Q statistic, calculated as the weighted sum of squared differences between individual SNP Wald estimates and the IVW estimate.

We adhered to the Strengthening the Reporting of Observational Studies in Epidemiology guidelines for the reporting of MR studies (STROBE-MR) [39] (Supplementary Table 1). All statistical analyses were conducted using R 4.5.1 (R Core Team, 2024).

## Results

### Genetic relevance and strength of BMI IV in African and European populations

A total of 428 genome-wide significant SNPs independently associated with BMI were identified in the meta-analyses of African and European populations. They included 363 (84.8%) that showed no evidence of heterogeneity (Cochran’s Q p-value > 0.1) and a direction of β-coefficients that was consistent across European and African populations. Therefore, 65 SNPs showed evidence of heterogeneity between African and European populations. Of these, 8 SNPs showed a stronger positive association of with BMI in Africans than Europeans, 2 SNPs showed a stronger positive association of increasing BMI in Europeans than Africans, and 55 SNPs associated positively with BMI in Europeans but negatively in Africans (Supplementary Figure 2). In our main analyses, we therefore used 363 cross-ancestry SNPs as IVs which were all strongly associated with BMI (median F-statistic of 42.1 (IQR: 34.8-60.2)), showing no evidence of weak instrument bias.

### Main two-sample MR and sensitivity analyses

The main IVW analyses showed a positive effect of BMI on HbA1c levels in both Africans (0.26 SD higher HbA1c per 1 SD BMI, 95% CI 0.07, 0.45) and Europeans (0.27 SD higher HbA1c per 1 SD BMI, 95% CI 0.22, 0.32) in UK Biobank, though the former was less precise than the European results, as expected given the smaller number of Africans than Europeans with HbA1c outcome data (Figure 1A). Sensitivity analyses using MR-Egger, weighted median and weighted mode were directionally consistent (Supplementary Table 2) and the p-values for the MR Egger intercept were 0.69 for Africans and 0.96 for Europeans.

**Figure 1:**
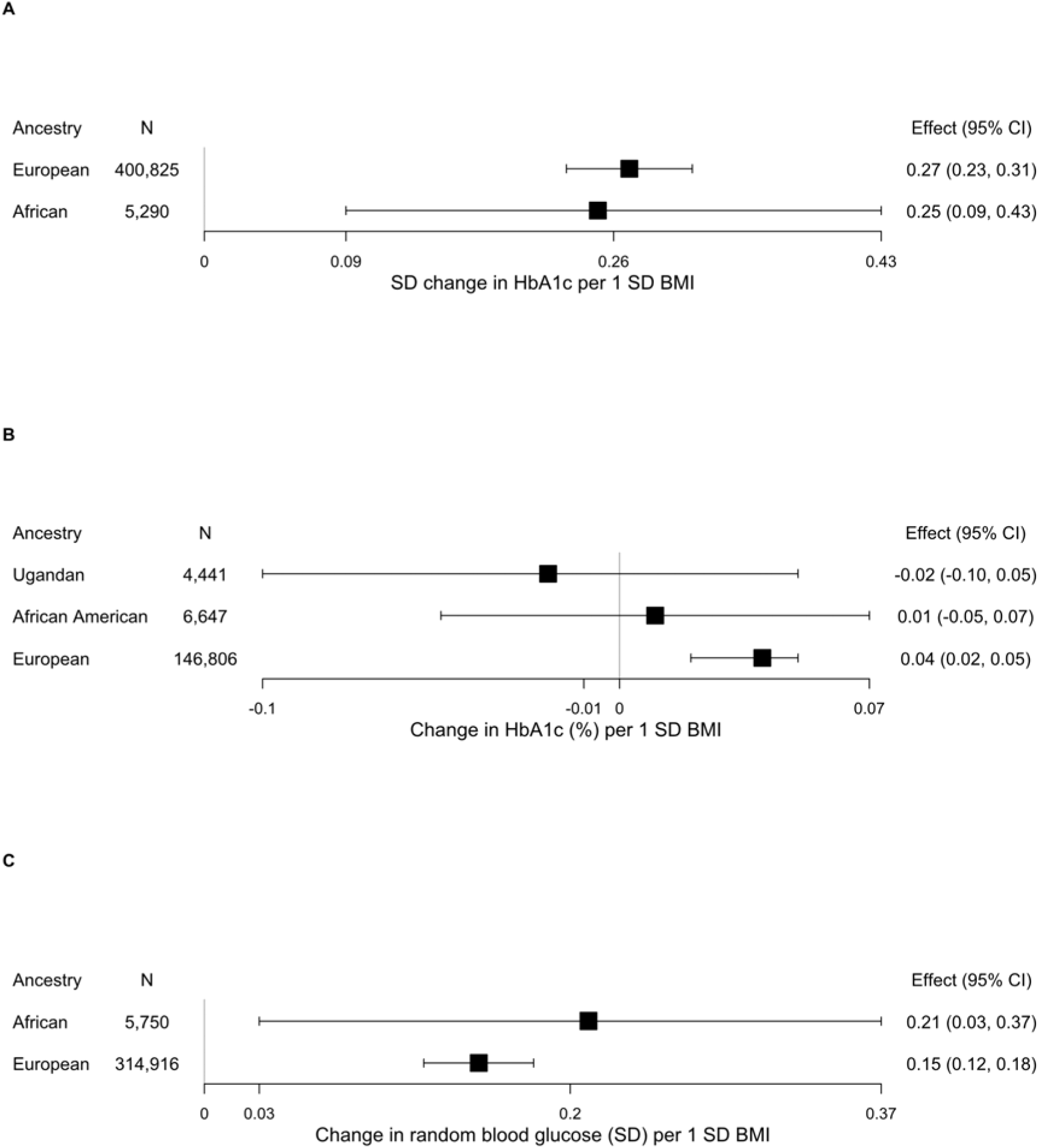
Mendelian randomization estimates of the effect between genetically predicted BMI and glycaemic traits for (A) HbA1c in UK Biobank participants, (B) HbA1c in the MAGIC cohort, and (C) random blood glucose in UK Biobank participants. Effect estimates are expressed in the outcome-specific units shown on each x-axis. Squares denote point estimates and horizontal lines represent 95% confidence intervals; the vertical reference line indicates no effect.

Similarly, when using data from the MAGIC consortia, the main IVW analyses provided evidence that BMI had a positive effect on HbA1c levels in African Americans (0.01% higher HbA1c per 1 SD BMI, 95% CI -0.05, 0.07) and Europeans (0.04 % higher HbA1c per 1 SD BMI, 95% CI 0.02, 0.05) but a negative effect in Ugandans (0.02% lower HbA1c per 1 SD BMI, 95% CI -0.10, 0.05) (Figure 1B). However, in both the African American and Ugandan populations the 95% CIs did span zero. Sensitivity analyses using MR-Egger, weighted median and weighted mode were directionally consistent and the p-values for the MR Egger intercept were 0.62 for Ugandans, 0.85 for African Americans, and 0.76 for Europeans (Supplementary Table 3).

The main IVW analyses for random glucose using UK Biobank data supported a positive causal effect of BMI in both Africans (0.21 SD higher mean glucose per 1 SD increase in BMI, 95% CI 0.03, 0.37) and Europeans (0.15 SD higher mean glucose per 1 SD increase in BMI, 95% CI 0.12, 0.18), though again the estimates were less precise in Africans than Europeans (Figure 1C). Sensitivity analyses using MR-Egger, weighted median and weighted mode were directionally consistent and the p-values for the MR Egger intercept were 0.49 for Africans and 0.82 for Europeans (Supplementary Table 4).

### Exploratory one sample analyses

In the one-sample MR, the BMI-weighted genetic risk had an F-statistic of 38.3. The BMI GRS was associated with a small increase in fasting glucose (mean difference 0.01 mmol/L for each 1 SD increase in BMI, 95% CI -0.01, 0.03). There was no evidence of heterogeneity between SNPs (Cochran’s Q = 397.7, p = 0.10; I^2^ = 9.0%) suggesting consistent SNP-specific estimates.

## Discussion

In this multi-ancestry study integrating data from AWI-Gen, GIANT, UK Biobank, and MAGIC, we investigated the transferability of BMI-associated variants across populations and the effects of genetically predicted BMI on glycaemic traits. We found that the majority of independent genome-wide significant BMI-associated SNPs exhibited concordant directions of effect in African and European populations, supporting a largely shared genetic architecture underlying BMI across ancestries. However, the effects of genetically predicted BMI on glycaemic traits varied across populations, highlighting the importance of including diverse ancestries in genetic epidemiological studies. Among SNPs that demonstrated heterogeneity, 55 showed opposite directions of effect between Africans and Europeans, suggesting the presence of population-specific genetic effects that may arise from differences in linkage disequilibrium structure, gene-gene interactions, or gene-environment interactions.

In our main two-sample MR using the UK Biobank, we showed that BMI had a positive causal effect on HbA1c in both African and European adults, and findings were consistent when conducting sensitivity analyses. The magnitude of effect was similar for both populations (0.26 SD higher HbA1c per 1 SD BMI in Africans and 0.27 SD higher HbA1c per 1 SD BMI in Europeans). In MAGIC, BMI had a positive causal effect on HbA1c among African Americans (0.01% higher per 1 SD BMI), although the 95% CI did span zero, and Europeans (0.04% higher per 1 SD BMI), consistent with previous MR evidence from European populations [11, 12]. In contrast, the effect estimate among Ugandans was in the opposite direction (0.02% lower HbA1c per 1 SD increase in BMI), and the 95% CIs spanned zero, providing limited evidence for a causal effect in this population. Evidence from non-European MR studies remains limited and inconsistent. For example, a Japanese MR study similarly reported no significant association between genetically predicted BMI and HbA1c (β = -0.040, 95% CI -0.081 to 0.002), suggesting that the causal effect of BMI on glycaemic regulation may vary across populations [40].

For random glucose, BMI demonstrated a positive causal effect in both Africans and Europeans. The effect estimate was numerically larger among Africans (0.21 SD increase in random glucose per 1 SD increase in BMI) compared to Europeans (0.15 SD increase in random glucose per 1 SD increase in BMI). Although evidence directly comparing ancestry-specific effects of adiposity on random glucose remains limited, ethnic differences in insulin sensitivity and β-cell function have been reported [41, 42]. Furthermore, recent evidence suggests that adiposity may be a key determinant of dysglycaemia and type 2 diabetes pathogenesis across populations despite differences in β-cell function [43]. Our sensitivity analysis on fasting glucose using one-sample MR in the AWI-Gen cohort yielded a directionally consistent effect estimate, although the 95% CIs did span zero.

Previous MR studies of the potential effects of BMI on glycaemic traits have been predominantly undertaken in European populations and largely explored effects on fasting glucose [12, 30, 44]. Our findings demonstrating positive effects of BMI on HbA1c and random glucose is consistent with those results, given the known correlations between these different glycaemic traits. Since previous MR studies have largely been restricted to European populations, our study expands this literature to include African populations.

There is evidence of differences in body fat distribution between African and European populations, with African individuals generally having lower visceral adipose tissue and ectopic fat deposition and greater gluteofemoral fat and lean mass than European individuals of the same BMI [45–47]. African populations have also been shown to exhibit differences in insulin physiology, characterised by low insulin sensitivity, increased insulin secretion and reduced hepatic insulin clearance which precipitate hyperinsulinaemia and higher glucose levels [45]. These differences in adiposity and insulin physiology at comparable levels of BM could plausibly contribute to differences between BMI and glycaemic traits across African and European populations [48, 49]. However, our results did not provide evidence for differences in the effects of BMI on the glycaemic traits examined between the two ancestry groups. There could be several reasons for this. Differences in body fat distribution between different ancestral groups may not be as marked or may not affect glycaemic traits as previously assumed. In support of this, MR studies using genetic variants as IVs for body composition have found similar associations between total adiposity and glycaemic traits and type 2 diabetes across different ancestries, including between African and European populations [50], which has some consistency with our findings. Furthermore, a previous study using data from the UK Biobank found no differences in visceral, liver, or intramuscular fat between European, South Asian and African populations [51]. However, the extent to which the effects of body adiposity on glycaemia are modified by contextual factors, such as the specific region from where a person originates (e.g., which part of Africa) and whether they are recent migrants or children of multiple generations of migrants is unclear. Importantly across these studies, Europeans are the dominant data-derived group and there remains a lack of genetic data and large cohort studies in African and other non-European groups. This is illustrated by the fact that we could not identify large GWAS of adipose traits in Africans living in Africa and whilst AWI-Gen is an important pioneering sub-Saharan African study, larger numbers of participants would be needed to undertake GWAS of adipose trait distribution in different geographic locations. It must be noted that body fat distribution, particularly central body fat, may be more strongly related to glycaemic traits than total adiposity, for which BMI is a proxy measure. Some MR studies have demonstrated causal effects of waist circumference and waist-to-hip ratio with glycaemic traits, with some evidence that central obesity may exert stronger effects than BMI on insulin- and glycaemic related traits [52, 53]. It would therefore be important for future MR studies to investigate the causal effects of different body fat depots on glycaemic traits across African and European populations.

### Strengths and Limitations

Key strengths of this study are its use of MR to compare effects of BMI on HbA1c and random glucose between African and European adults and use of a multi-ancestry GWAS meta-analysis to derive equally strong genetic instruments for comparative MR analyses in African and European populations. Strengths also include undertaking multiple sensitivity analyses and an exploratory one sample MR analysis in an African cohort.

Our study had several limitations. First, the African data for HbA1c and random glucose GWAS was based on substantially smaller numbers of participants than the corresponding European GWAS. Whilst our results suggest broadly similar increases in HbA1c and random glucose with higher BMI, the imprecision in the results from the African population means that the data should be interpreted with caution. Secondly, in the European MR analysis, our study design included overlapping samples of exposure and outcome as UK Biobank participants were included in the GIANT BMI GWAS which could lead to the overfitting of the causal estimate. Thirdly, as with any MR study we are reliant on the availability of relevant and well conducted GWAS. Ideally, we would have explored effects on additional glycaemic traits, such as fasting glucose, post-load glucose and fasting insulin, but we were aware that even in European populations GWAS of these outcomes commonly adjust for BMI and other heritable traits and studies in Africans for these outcomes are small or absent. Fourthly, we were only able to study the relationship of BMI with glycaemic traits but inter-population differences in glycaemia may be influenced more by variation in body fat distribution. However, GWAS for these anthropometric measures are less common than for BMI, especially in African populations.

## Conclusion

This study provides evidence supporting a causal role of higher BMI in increasing HbA1c and glucose levels among African and European ancestry populations. The method we used to obtain strong genetic BMI IVs combining data from GWAS in African and European studies to apply in both groups may have value for other MR studies interested in exploring differences in effects between different population groups. This study also highlights the need for much larger GWAS and increased investment in African, and other non-European populations, to improve our understanding of global health and reduce global inequalities.

## Data Availability

All data produced in the present work are contained in the manuscript

https://www.ebi.ac.uk/gwas/summary-statistics

https://gwas.mrcieu.ac.uk/datasets/

## Abbreviations

*AADM*: Africa America Diabetes Mellitus
*AWI-Gen*: Africa-Wits International Network for the Demographic Evaluation of Populations and Their Health (INDEPTH) Partnership for Genomic Studies
*BMI*: body mass index
*DCC*: Diabetes Case Control
*DDS*: Durban Diabetes Study
GIANT: Genetic Investigation of ANthropometric Traits
*GWAS*: Genome-wide association study/studies
*HbA1c*: Glycated haemoglobin
*IEU OpenGWAS*: Integrative Epidemiology Unit OpenGWAS database
*IV*: Instrumental variables
IVW: Inverse-variance weighted
*LD*: Linkage disequilibrium
*MR*: Mendelian randomization
*SNPs*: single nucleotide polymorphisms

## Ethics declarations

## Acknowledgements

We thank the AWI-Gen participants, study centre leadership, study coordinators, laboratory scientists, data managers and field teams at the study centres of this work. Computing was performed on the Wits Core Research Cluster at the University of Witwatersrand. We thank the participants and researchers from GIANT, MAGIC and UK Biobank, who contributed data.

## Funding

The AWI-Gen study was funded by the National Human Genome Research Institute, Office of the Director, the Eunice Kennedy Shriver National Institute of Child Health & Human Development, the National Institute of Environmental Health Sciences, the Office of AIDS Research, and the National Institute of Diabetes and Digestive and Kidney Diseases of the National Institutes of Health (grant number U54HG006938 and its supplements), as part of the H3Africa Consortium and the Department of Science and Innovation, South Africa (grant number DST/CON 0056/2014). RPC was funded by the 2023 Meade Travel Grants in Epidemiology from the Medical Research UK (MEG2\165), which supported a research visit to the University of Bristol for training in Mendelian randomization methods. RPC was also supported by the Sydney Brenner Institute for Molecular Bioscience and the Department of Science and Technology and Innovation-National Research Foundation (DSTI-NRF) Centre of Excellence in Human Development at the University of the Witwatersrand, Johannesburg. None of the funding sources were involved in any parts of the study nor in writing the manuscript and the decision to submit it for publication. DAL’s contribution to this study is supported by the UK Medical Research Council (MC_UU_00032/05) and the British Heart Foundation (CH/F/20/90003). AGS, AHWC and DAL work in a unit supported by the University of Bristol and UK Medical Research Council (MC_UU_00032/5) and the British Heart Foundation (AA/18/1/34219). AGS and DAL are supported by STAGE that has received funding from the European Union’s Horizon Europe Research and Innovation Programme under grant agreement number 101137146 (via UKRI grant number 10099041).

None of the funders influenced study analyses or interpretation of results.

The views expressed in this paper are those of the authors and not necessarily any funding organisation or people acknowledged.

## Authors’ relationships and activities

DAL reports participation in (i) research funded by the Diabetes UK and the European Research Council that is not related to this paper (funding related to this paper is declared in the funding section), (ii) being on the UK Biobank advisory group (no funding), and (iii) being a member of the Lancet Obstetrics, Gynaecology and women’s health journal (no funding). The remaining authors declare that there are no relationships or activities that might bias, or be perceived to bias, their work.

## Contribution statement

DAL, AHWC, AGS, RPC, MR, NJC designed this study

RPC, DAL, AHWC, and AGS wrote the analysis plan

MR and NJC designed, managed data collection and manage the AWI-Gen cohort

RPC, AHWC, and DAL wrote the initial analysis plan with further input from AGS

DS undertook the AWI-Gen BMI GWAS

RPC undertook the Mendelian Randomization analyses and interpretation with guidance from JTB, DAL, AHWC and AGS

RPC wrote the first draft, with input from DAL

RPC, JTB and MR had full access to all the data in the study and take responsibility for the integrity of the data and the accuracy of the data analysis.

All authors provided critical feedback and approved the final manuscript.

## Data availability

The GWAS datasets used in the current study were derived from the IEU OpenGWAS (https://gwas.mrcieu.ac.uk/datasets/) and the GWAS Catalog (https://www.ebi.ac.uk/gwas/summary-statistics). From IEU OpenGWAS, summary statistics used were: ukb-d-30740_irnt.vcf.gz, ukb-e-30740_AFR.vcf.gz and biomarkers-30750-both_sexes-irnt.tsv.bgz. From the GWAS catalog, summary statistics accessed included: GCST90002242, GCST90002244, and GCST90002248. De-identified individual participant data from the AWI-Gen study are available from the European Genome-Phenome Archive (accession number EGA00001002482). The Human Heredity and Health in Africa (H3Africa) Data and Biospecimen Access Committee will review requests for the AWI-Gen phenotype and genotype datasets.

## Code availability

Source code implementing the methods described in the paper are publicly available at https://github.com/rayltonc/BMI-and-Glycaemic-Traits-Mendelian-Randomization.

## Notes

### Author Declarations

Ethical approval for the recruitment of participants in the AWI-Gen study was obtained from the Human Research Ethics Committee (Medical) of the University of the Witwatersrand (M121029 and M2210108). All participants provided written informed consent following community engagement and individual information sharing. The MR analyses used publicly available GWAS summary statistics from studies that had received ethical approval from their respective institutions and obtained informed consent from participants. Only anonymised summary-level data were accessed; therefore, no additional ethical approval was required for these analyses.

