## Supplementary text for "Causal effects of body mass index on glycaemic traits in African and European populations: a Mendelian randomization study"

On May 10, 2026, we searched the GWAS catalog [17] IEU OpenGWAS [18], and PubMed for large GWAS of glycaemic traits to be used as outcomes in Mendelian randomization analyses. Searches incorporated terms related to glycaemic traits (fasting glucose, random glucose, insulin, HbA1c, glycated haemoglobin, and 2-hour glucose), GWAS methodology (GWAS, genome-wide association study, meta-analysis, and summary statistics), and ancestry (European and African). Consortium-specific repositories were also searched for eligible studies available up to May 2026.

We identified the Meta-Analyses of Glucose and Insulin-related traits Consortium (MAGIC) and UK Biobank as sources of relevant data with both African and European-specific summary data. From the MAGIC consortia, summary data was available for fasting glucose, 2-hour glucose, glycated haemoglobin (HbA1c), and fasting insulin data. However, only HbA1c summary data without BMI adjustment was available from MAGIC [31]. From UK Biobank, only HbA1c and random glucose summary data were available, without BMI adjustment [33].
