## Supplementary Table 2 for "Causal effects of body mass index on glycaemic traits in African and European populations: a Mendelian randomization study"

**Supplementary Table 2:** Mendelian randomization causal effect of BMI on HbA1c GWAS from the UK Biobank

|  | European (n=400,825) | | | African (n=5,290) | | |
| --- | --- | --- | --- | --- | --- | --- |
| Method | SD HbA1c per 1 SD BMI increase | 95% CI | *p* value | SD HbA1c per 1 SD BMI increase | 95% CI | *p* value |
| IVW | 0.27 | 0.22, 0.32 | 3.16 x 10^-26^ | 0.26 | 0.07, 0.45 | 6.67 x 10^-3^ |
| MR-Egger  Slope | 0.27 | 0.10, 0.43 | 1.45 x 10^-3^ | 0.26 | 0.07, 0.45 | 7.10 x 10^-3^ |
| MR-Egger intercept | <-0.01 | <-0.01, <0.01 | 0.96 | <-0.01 | <-0.01, <0.01 | 0.69 |
| Weighted median | 0.29 | 0.26, 0.36 | 4.40 x 10^-30^ | 0.23 | -0.07, 0.52 | 0.12 |
| Weighted mode | 0.36 | 0.29, 0.43 | 6.80 x 10^-25^ | 0.11 | -0.89, 1.12 | 0.83 |
