## Supplementary Table 3 for "Causal effects of body mass index on glycaemic traits in African and European populations: a Mendelian randomization study"

**Supplementary Table 3:** Mendelian randomization causal effect of BMI on HbA1c GWAS from the MAGIC consortium

| MAGIC | Ugandans (n=4,441) | | | African Americans (n=6,647) | | | European (n=146,806) | | |
| --- | --- | --- | --- | --- | --- | --- | --- | --- | --- |
|  | % HbA1c per 1 SD BMI | 95% CI | *p* value | % HbA1c per 1 SD BMI | 95% CI | *p* value | % HbA1c per 1 SD BMI | 95% CI | *p* value |
| IVW | -0.02 | -0.10, 0.05 | 0.53 | 0.01 | -0.05, 0.07 | 0.77 | 0.04 | 0.02, 0.05 | 1.95 x 10^-8^ |
| MR-Egger  Slope | -0.02 | -0.10, 0.05 | 0.54 | 0.01 | -0.05, 0.07 | 0.80 | 0.04 | 0.02, 0.05 | 2.17 x 10^-8^ |
| MR-Egger intercept | 3.46 x 10^-4^ | -1.03 x 10^-3^, 1.72 x 10^-3^ | 0.62 | -1.05 x 10^-4^ | -1.19 x 10^-3^, 9.76 x 10^-4^ | 0.85 | 3.49 x 10^-5^ | -1.90 x 10^-4^, 2.60 x 10^-4^ | 0.76 |
| Weighted median | 0.03 | -0.12, 0.12 | 0.93 | -0.02 | -0.02, 0.08 | 0.61 | 0.04 | 0.03, 0.06 | 2.06 x 10^-9^ |
| Weighted mode | 0.05 | -0.31, 0.41 | 0.79 | -0.05 | -0.34, 0.71 | 0.71 | 0.07 | 0.03, 0.11 | 1.44 x 10^-4^ |
