## Supplementary Table 4 for "Causal effects of body mass index on glycaemic traits in African and European populations: a Mendelian randomization study"

**Supplementary Table 4:** Mendelian randomization causal effect of BMI on random glucose GWAS from UK Biobank

|  | African (n=5,750) | | | European (n=314,916) | | |
| --- | --- | --- | --- | --- | --- | --- |
|  | 1 SD random blood glucose per 1 SD BMI | 95% CI | *p* value | 1 SD random blood glucose per 1 SD BMI | 95% CI | *p* value |
| IVW | 0.21 | 0.03, 0.37 | 6.67 x 10^-3^ | 0.15 | 0.12, 0.18 | 1.04 x 10^-20^ |
| MR-Egger  Slope | 0.22 | 0.07, 0.38 | 5.59 x 10^-3^ | 0.13 | 0.06, 0.21 | 1.04 x 10^-3^ |
| MR-Egger intercept | -9.57 x 10^-4^ | -3.68 x 10^-3^, 1.77 x 10^-3^ | 0.49 | 1.61 x 10^-4^ | -1.27 x 10^-3^,  1.59 x 10^-3^ | 0.82 |
| Weighted median | 0.09 | -0.07, 0.26 | 0.16 | 0.17 | 0.13, 0.18 | 1.54 x 10^-39^ |
| Weighted mode | 0.02 | -0.80, 0.85 | 0.96 | 0.18 | 0.12, 0.25 | 1.47 x 10^-8^ |
