## Supplementary Figure 1 for "Causal effects of body mass index on glycaemic traits in African and European populations: a Mendelian randomization study"

SNPs imputation

11,702 participants 2,267,346 SNPs

SNPs exclusions

Missingness >0.05 Hardy-Weinberg equilibrium <0.0001 MAF <0.01, Info score <0.6 Non-autosomes, Duplicates Mismatches to GRCh37 build

Sample exclusions

Missingness >0.05 Heterozygosity (<0.15 and >0.34) Duplicates (PIHAT >0.9) Sex inconsistencies

Final genotype data 10,603 participants 1,733,121 SNPs

Quality control 127 participants removed (67 PCA outliers, 58 missing BMI, 2 extreme BMI values)

10,476 participants for BMI GWAS 13,980,000 SNPs

**Supplementary Figure 1:** Flowchart showing pre- and post-imputation quality control on genotyped data from the AWI-Gen study population.
