## Supplementary Figure 2 for "Causal effects of body mass index on glycaemic traits in African and European populations: a Mendelian randomization study"

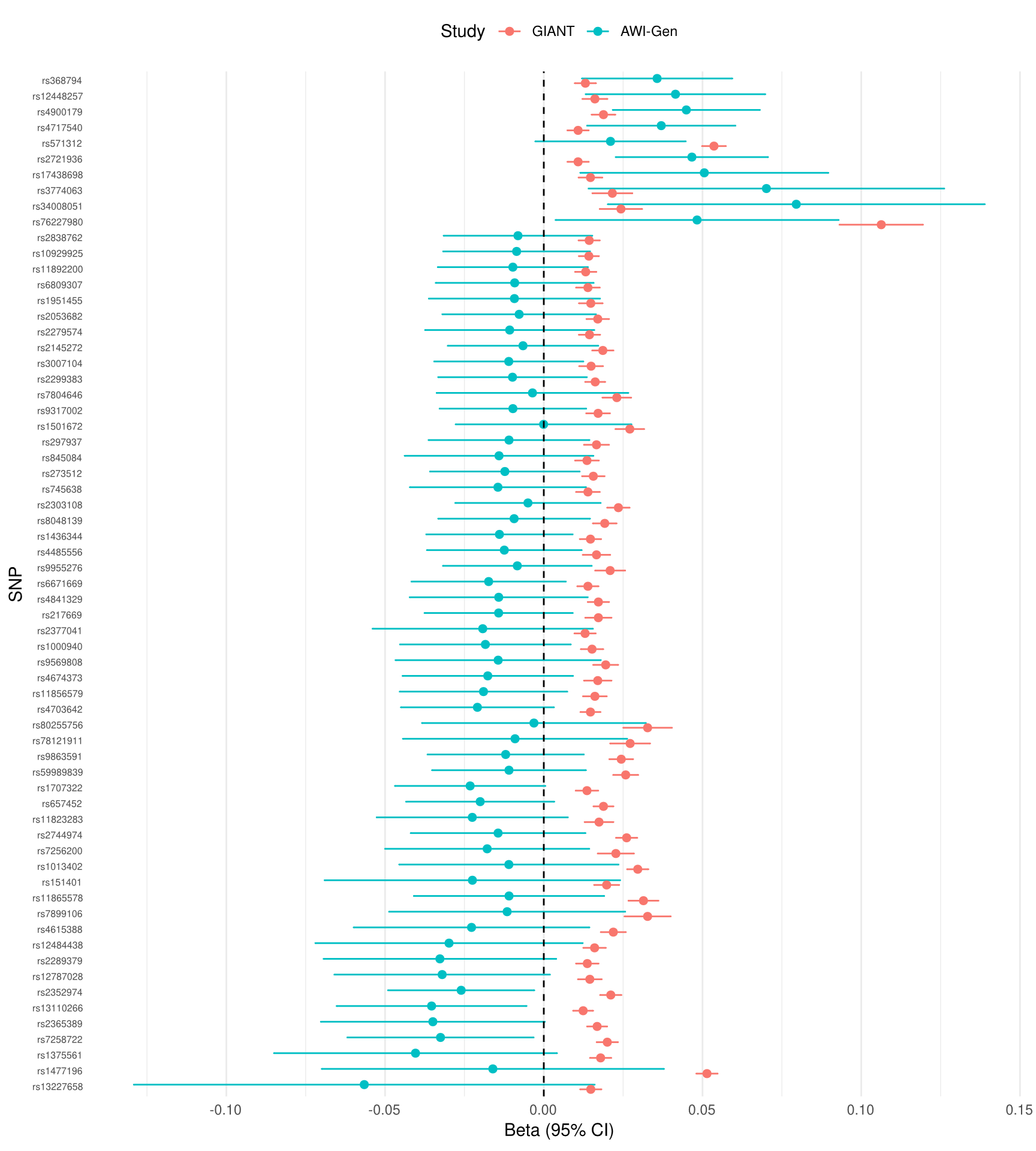

**Supplementary Figure 2:** Forest plot of heterogeneity between 65 AWI‑GEN (African ancestry) and GIANT (European ancestry) BMI genetic instruments (SNPs) that showed evidence of heterogeneity (Cochran’s Q p‑value <0.1).
